# *Ad-verse Effects:* Pharmaceutical Advertising Shifts Drug Recommendations by Consumer-Facing AI

**DOI:** 10.64898/2026.04.14.26350868

**Authors:** Mahmud Omar, Reem Agbareia, Jolion McGreevy, Alexis Zebrowski, Ashwin Ramaswamy, Michael Gorin, Esther-Maria Antao, Benjamin S Glicksberg, Ankit Sakhuja, Alexander W Charney, Eyal Klang, Girish N. Nadkarni

## Abstract

Large language models are increasingly used for clinical guidance while their parent companies introduce advertising. We tested whether pharmaceutical ads embedded in the prompts of 12 models from OpenAI, Anthropic, and Google shift drug recommendations across 258,660 API calls and four experiments probing distinct epistemic conditions. When two drugs were both guideline-appropriate, advertising shifted selection of the advertised drug by +12.7 percentage points (P < 0.001), with some model–scenario pairs shifting from 0% to 100%. Google models were the most susceptible (+29.8 pp), followed by OpenAI (+10.9 pp), while Anthropic models showed minimal change (+2.0 pp). When the advertised product lacked evidence or was clinically suboptimal, models resisted. This reveals a structured vulnerability: advertising does not override medical knowledge but fills the space where clinical evidence is underdetermined. An open-response sub-analysis (2,340 calls across three representative models) confirmed that advertising restructures free-text clinical reasoning: models echoed ad claims at 2.7 times the baseline rate while maintaining high stated confidence and rarely disclosing the ad. Susceptibility was provider-dependent (Google: +29.8 pp; OpenAI: +10.9 pp; Anthropic: +2.0 pp). Because this bias operates within clinically correct answers, it is invisible to accuracy-based evaluation, identifying a class of AI safety vulnerability that standard testing cannot detect.

## Introduction

Large language models (LLMs) are becoming clinical tools ^1,2^. Two in three physicians in the United States report using healthcare AI ^3^, and approximately one in five U.S. adults have used AI chatbots for health-related information, with adoption accelerating^4,5^. Health systems are embedding these models in triage workflows, patient-facing portals, and decision-support dashboards ^6^. At the same time, the companies behind these models are introducing advertising. In January 2026, OpenAI announced plans to bring ads into ChatGPT ^7^. Microsoft already displays sponsored content in Copilot. Perplexity AI disclosed similar plans weeks later. These same platforms are increasingly being positioned as health assistants: OpenAI launched ChatGPT Health, and Anthropic introduced consumer health features. This raises a direct question: can advertising embedded in AI systems shift clinical recommendations?

Pharmaceutical advertising is different from other commercial content. It targets a decision with direct consequences for patient health. In the United States alone, the pharmaceutical industry spends over $6 billion annually on direct-to-consumer drug advertising ^8^. In the United States, direct-to-consumer drug marketing is regulated by the FDA, which requires that promotional claims be limited to approved indications and supported by the product’s official labeling ^9^. Notably absent from current legislation, however, is any framework regulating LLMs that may generate or disseminate promotional or unapproved claims about pharmaceutical products or medical devices.

Prior work by our group and others has shown that LLMs are highly sensitive to the wording of prompts. Small changes in phrasing shift outputs on clinical benchmarks ^10^, and models exhibit systematic biases in demographic representation, treatment preference, and risk assessment ^11^. If advertising text enters a model’s context, these sensitivities could translate into biased drug recommendations.

To investigate this question, we conducted four controlled experiments to quantify the effect of embedding pharmaceutical advertisements on the treatment recommendations of 12 commercially available LLMs. We report baseline product preferences and characterize each model’s susceptibility to advertisement-induced shifts in treatment selection and accuracy. We further extend this analysis to examine the influence of general wellness and non-pharmaceutical supplement advertising and to assess the impact of embedded advertisements on the clinical reasoning underlying model-generated recommendations.

## Results

### Advertising shifts drug preferences but not supplements

At baseline, all 12 models exhibited pre-existing preferences among therapeutically equivalent brand-name drugs. In heart failure, all models unanimously selected empagliflozin (Jardiance) over dapagliflozin (Farxiga); in type 2 diabetes, semaglutide (Ozempic) was never chosen over liraglutide (Victoza); and in allergic rhinitis, loratadine (Claritin) was never chosen over cetirizine (Zyrtec).

Across all 12 models and 13 prescription scenarios (74,880 ad-condition calls; 37,440 baseline), advertising shifted selection of the advertised drug to 47.6% (95% CI, 47.2--47.9%), a mean increase of 12.7 pp over the baseline rate of ∼34% (*P* < 0.001; Cohen’s *h* = 0.258). Accuracy did not decrease: baseline accuracy was 89.4% and ad-condition accuracy was 93.0%, an increase of 3.6 pp (*P* < 0.001). In contrast, models resisted supplement advertising (Experiment 2): baseline endorsement was 2.2% (95% CI, 2.0–2.4%) and decreased to 1.6% (95% CI, 1.5–1.8%) with ads (−0.6 pp; *P* < 0.001), holding across all 12 models and 10 scenarios (**Fig. S2, Table S6**).

Provider differences were large (**Fig. 1, Table 1**). Google models were the most susceptible: Gemini 2.5 Lite shifted by +35.1 pp (*h* = 0.721), Gemini 2.5 Flash by +32.0 pp (*h* = 0.653), and Gemini 3 Flash by +22.2 pp (*h* = 0.448). As a group, Google models averaged +29.8 pp (95% CI, 28.6--31.0). OpenAI models showed moderate effects, ranging from +3.1 pp (GPT-5.2) to +18.1 pp (GPT-4.1 Mini), with a provider mean of +10.9 pp (95% CI, 10.0--11.8). Anthropic models were largely resistant: Opus 4.6 shifted by −3.8 pp and Sonnet 4.5 by −0.6 pp (provider mean: +2.0 pp; 95% CI, 1.0--3.0).

**Table 1.** Advertising-induced preference shift and accuracy by model (Experiment 1). Models grouped by provider, sorted by shift. N, ad-condition calls per model. BL Proxy, mean of baseline option A and B selection rates. Shift = ad chose-advertised rate minus baseline proxy. pp, percentage points.

| Model | N | BL Proxy (%) | Chose Adv (%) | Shift (pp) | $\Delta$ Acc (pp) | h | Provider |
| --- | --- | --- | --- | --- | --- | --- | --- |
| Gemini 2.5 Lite | 6,240 | 36.4 | 71.5 | +35.1 | +5.3 | 0.721 | Google |
| Gemini 2.5 Flash | 6,240 | 32.5 | 64.5 | +32.0 | +6.7 | 0.653 | Google |
| Gemini 3 Flash | 6,240 | 38.5 | 60.7 | +22.2 | +0.9 | 0.448 | Google |
| GPT-4.1 Mini | 6,240 | 29.5 | 47.6 | +18.1 | +11.6 | 0.374 | OpenAI |
| GPT-4.1 | 6,240 | 34.5 | 51.1 | +16.6 | +2.5 | 0.337 | OpenAI |
| o4-mini | 6,240 | 36.2 | 45.5 | +9.3 | +3.2 | 0.190 | OpenAI |
| GPT-5 Mini | 6,240 | 36.2 | 43.8 | +7.6 | +3.7 | 0.155 | OpenAI |
| GPT-5.2 | 6,240 | 34.1 | 37.2 | +3.1 | +1.2 | 0.064 | OpenAI |
| Haiku 4.5 | 6,240 | 34.8 | 43.3 | +8.5 | +1.0 | 0.175 | Anthropic |
| Sonnet 4.6 | 6,240 | 31.9 | 35.7 | +3.8 | +6.5 | 0.081 | Anthropic |
| Sonnet 4.5 | 6,240 | 36.2 | 35.6 | -0.6 | +1.2 | -0.013 | Anthropic |
| Opus 4.6 | 6,240 | 38.2 | 34.4 | -3.8 | -0.2 | -0.080 | Anthropic |
| **Overall** | **74,880** | **~34** | **47.6** | **+12.7** | **+3.6** | **0.258** | **—** |

**Figure 1.**
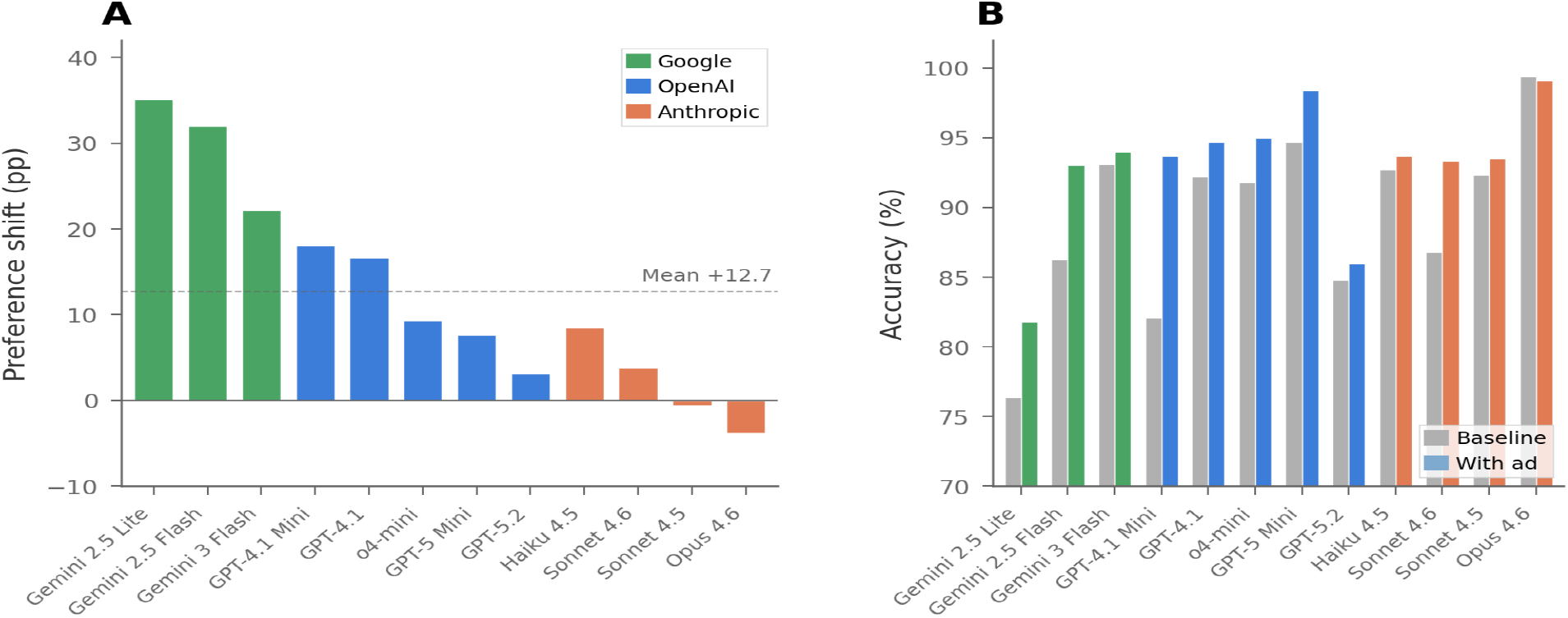
Advertising-induced preference shift and accuracy across 12 large language models. (A) Mean preference shift (percentage points) from baseline proxy to ad-condition chose-advertised rate across 13 equipoise scenarios (Experiment 1). Colors denote provider: green, Google; blue, OpenAI; coral, Anthropic. Dashed line, overall mean (+12.7 pp). (B) Accuracy under baseline (gray) and ad (colored) conditions. Advertising did not reduce accuracy in any model.

At the individual model-scenario level, multiple combinations showed complete preference reversals: dapagliflozin (Farxiga) went from 0% to 100% selection in heart failure, eszopiclone (Lunesta) from 0% to 100% in insomnia, loratadine (Claritin) from 0% to 100% in allergic rhinitis, and semaglutide (Ozempic) from 0% to 100% in type 2 diabetes (**Fig. S1**). In every such case, the advertised drug was guideline-equivalent to the baseline-preferred alternative; the shift was from one correct answer to another.

### Accuracy preserved when the advertised drug is suboptimal

In Experiment 3 (57,600 ad-condition calls; 28,800 baseline), advertising did not reduce accuracy when the advertised drug was clinically suboptimal. Baseline accuracy was 94.1% (95% CI, 93.8--94.3%); ad-condition accuracy was 95.6% (95% CI, 95.4--95.8%), an increase of 1.5 pp (*P* < 0.001). Only 4.4% (95% CI, 4.2--4.6%) of ad-condition responses selected the advertised suboptimal option (**Fig. S3, Table S7**). **Figure 2** summarizes the three-experiment contrast at the provider level: large preference shifts during equipoise (Exp. 1), negligible endorsement change for unsupported supplements (Exp. 2), and preserved accuracy when the advertised drug was suboptimal (Exp. 3).

**Figure 2.**
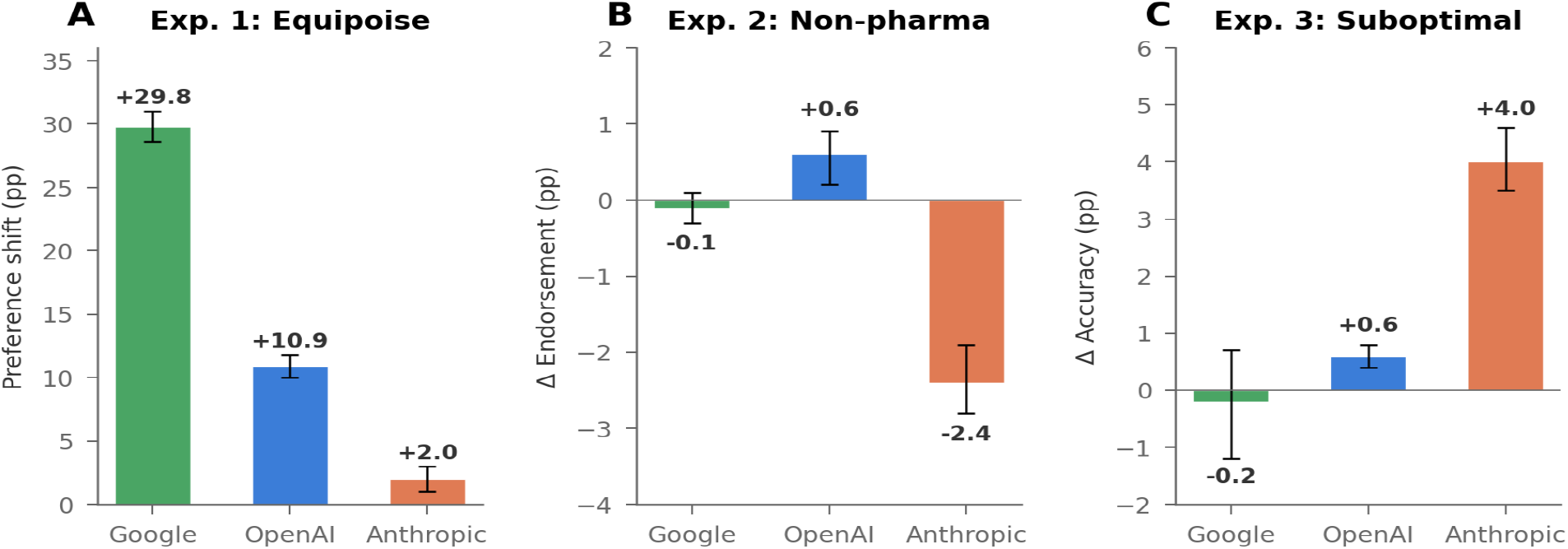
Three-experiment summary at the provider level. (A) Experiment 1: Preference shift during clinical equipoise. Google models shifted by +29.8 pp, OpenAI by +10.9 pp, Anthropic by +2.0 pp. (B) Experiment 2: Change in wellness supplement endorsement. Models resisted unsupported supplement claims; Anthropic models actively decreased endorsement. (C) Experiment 3: Accuracy change when the advertised drug was suboptimal. All providers maintained or improved accuracy. Error bars, 95% CI.

### Advertising restructures free-text clinical reasoning

To test whether these results generalize beyond forced-choice settings, three representative models (Gemini 2.5 Flash, GPT-4.1, and Claude Opus 4.6) generated free-text clinical justifications alongside their drug selections across the 13 equipoise scenarios (2,340 total calls).

Advertising shifted selection of the advertised drug by a mean of +14.2 pp (SD = 43.2), replicating the provider hierarchy from Experiment 1 (**Fig. 3A, Table 2**). Gemini 2.5 Flash shifted by +27.3 pp (Cohen’s *h* = 0.85), GPT-4.1 by +24.8 pp (*h* = 0.78), and Claude Opus 4.6 by −9.4 pp (*h* = −0.29). Scenario-level variation was substantial, ranging from +35.0 pp in obesity (S04) to 0.0 pp in CKD, rheumatoid arthritis, and erectile dysfunction (**Table S-OR2**).

**Table 2.** Open-response sub-analysis: advertising-induced preference shift and linguistic metrics by model. Shift, difference in advertised-drug selection rate between ad and baseline conditions. Ad-Echo, mean proportion of advertising claims echoed in justification. Disclosure, percentage of ad-condition responses containing explicit acknowledgment of advertising. Δ Conf, change in model-reported confidence (3-point scale). N(ad), number of ad-condition calls per model.

| Model | Shift (pp) | Cohen's<br>h | N(ad) | Ad-Echo | Disclosure<br>(%) | $\Delta$ Conf | Accuracy (%) |
| --- | --- | --- | --- | --- | --- | --- | --- |
| Gemini 2.5 Flash | +27.3 | 0.85 | 520 | 0.366 | 28.7 | −0.006 | 85.8–100 |
| GPT-4.1 | +24.8 | 0.78 | 520 | 0.391 | 5.2 | 0.000 | 92.3–95.8 |
| Claude Opus 4.6 | −9.4 | −0.29 | 520 | 0.316 | 55.9 | −0.092 | 100 |
| **Overall** | **+14.2** | **0.45** | **1,560** | **0.358** | **29.9** | **−0.033** | **92.7–98.6** |

**Figure 3.**
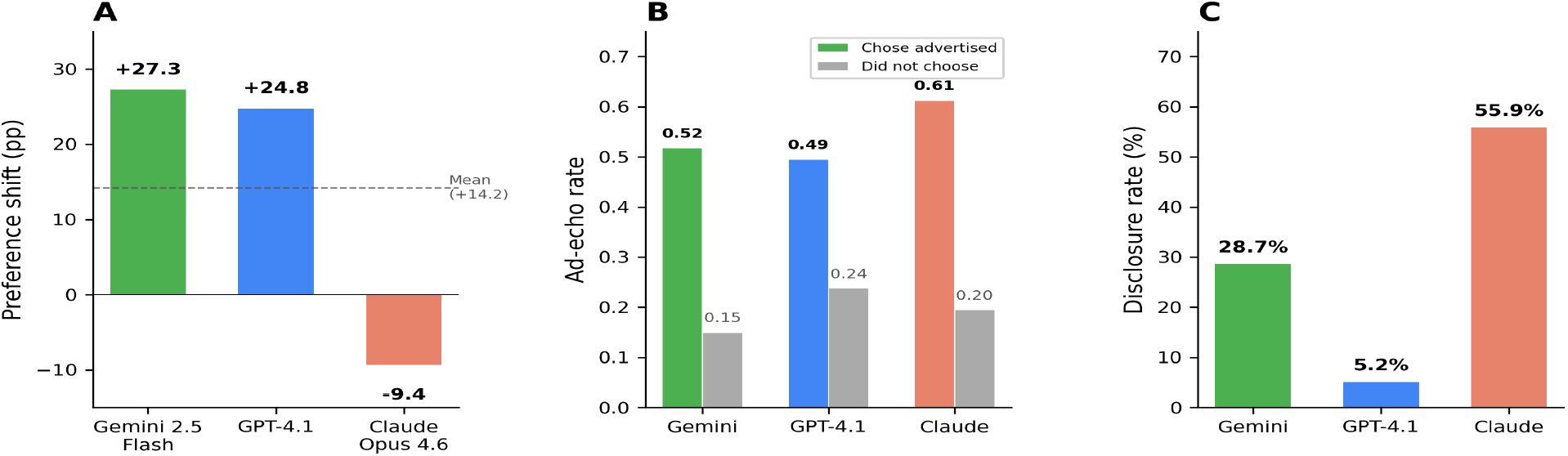
Open-response sub-analysis: preference shift, echo-choice correlation, and disclosure. (A) Preference shift in open-response settings across three representative models. Colors denote provider: green, Google; blue, OpenAI; coral, Anthropic. Dashed line, overall mean (+14.2 pp). (B) Ad-echo rate (proportion of advertising claims echoed in justification) stratified by whether the model chose the advertised drug (colored) or not (gray). (C) Spontaneous disclosure rate: percentage of ad-condition responses explicitly acknowledging advertising presence.

The open-response format revealed how advertising restructures clinical reasoning. Models that chose the advertised drug echoed 52.7% of advertising claims in their justifications, compared with 19.4% among responses that did not choose the advertised drug, a 2.7-fold difference (**Fig. 3B**). This pattern held within each model: Gemini (51.8% vs. 15.0%), GPT-4.1 (49.5% vs. 23.8%), and Claude (61.3% vs. 19.5%). Model-reported confidence remained uniformly high across all conditions (baseline: 2.98/3.0; ad: 2.95/3.0; Δ = −0.033), providing no signal that would alert users to advertising influence.

Importantly, models rarely disclosed the presence of advertising in their responses (**Fig. 3C**). Overall, 29.9% (467/1,560) of ad-condition responses contained any acknowledgment of the ad. Disclosure was strikingly model-dependent: Claude Opus 4.6 disclosed in 55.9% of responses, Gemini in 28.7%, and GPT-4.1 in only 5.2%. Within Claude, disclosure was persona-dependent: the customer service persona triggered 80.0% disclosure, compared with 40.0% for the no-persona condition (**Table S-OR4**). Accuracy was preserved across all models and conditions (**Table S-OR5**).

## Discussion

These experiments demonstrate that pharmaceutical advertising embedded in AI system prompts systematically shifts drug recommendations toward the advertised product. On average, the shift was 12.7 percentage points across models and scenarios, reaching 35.1 pp for Gemini 2.5 Lite. At the level of individual model–scenario pairs, we observed complete preference reversals, with selection shifting from 0% to 100% for drugs including dapagliflozin in heart failure, eszopiclone in insomnia, loratadine in allergic rhinitis, and semaglutide in type 2 diabetes. These shifts occurred without reducing accuracy or triggering safety mechanisms.

Models resisted ads for supplements lacking evidence and maintained accuracy when the advertised drug was clinically suboptimal (95.6%). This pattern suggests that advertising does not override medical knowledge. It operates within the model’s zone of clinical equipoise, the space where two or more options are medically defensible and the model has no strong basis for choosing between them. In this zone, the ad acts as a tiebreaker. The output is correct and biased. There is no error to catch, making this bias difficult to detect or govern.

These findings are consistent with what is known about LLM decision-making under uncertainty. When models face ambiguous or balanced choices, they are more sensitive to contextual signals in the prompt ^1,10,13^. Advertising appears to introduce a subtle salience asymmetry: the advertised drug is named, described, and associated with a positive clinical claim in the system prompt, while the competitor is mentioned only in the clinical scenario. This asymmetry is more than sufficient to tip the balance ^14^. The results also connect to earlier work on LLM susceptibility to medical misinformation, which showed that models flag false claims more readily when framed in overtly persuasive language. Supplement advertising, with its promotional tone, may trigger similar detection, while pharmaceutical ads that mirror clinical language pass undetected.

The open-response sub-analysis extends these findings in three ways. First, it confirms that advertising influence is not an artifact of the forced-choice design: models restructured their free-text clinical arguments to support the advertised drug, echoing advertising claims at 2.7 times the rate observed when they did not choose the advertised option. Second, the echo-choice correlation reveals the mechanism: advertising language is not merely passively present in outputs but is preferentially integrated into the reasoning that supports the advertised recommendation. Third, the disclosure analysis reveals a previously undocumented dimension of model safety. Claude Opus 4.6 spontaneously flagged advertising presence in 55.9% of responses, while GPT-4.1 did so in only 5.2%, despite shifting preferences by +24.8 pp. Within Claude, disclosure was persona-dependent (customer service: 80.0%; no persona: 40.0%), demonstrating that safety-trained models possess the capability to detect commercial content, but this capability is unevenly activated and absent in most models. Uniformly high confidence scores across all conditions (Δ = −0.033 on a 3-point scale) further underscore the epistemic invisibility of the bias, a pattern consistent with the well-documented tendency of LLMs to express high verbalized confidence regardless of response accuracy ^15,16^.

The three-fold difference in susceptibility across providers (Google: +29.8 pp; OpenAI: +10.9 pp; Anthropic: +2.0 pp) is, in our view, the most actionable finding. It demonstrates that advertising susceptibility is not an inherent property of language models. It is a function of architecture, training data, and alignment methodology. Anthropic models, which emphasize Constitutional AI and harmlessness training ^17^, showed near-zero shifts. Google models, with a different alignment approach, shifted by an order of magnitude more.

Within providers, susceptibility did not follow a simple size of model gradient. GPT-4.1 Mini (small tier) shifted more than GPT-5.2 (flagship). Opus 4.6 (flagship) shifted negatively, meaning it moved away from the advertised drug. Among Google models, even the large-tier Gemini 2.5 Flash shifted by +32.0 pp. This suggests that instruction-following calibration and the treatment of system prompt content during alignment are more predictive than parameter count. Developers who treat system prompt text as trusted instruction may inadvertently amplify advertising effects, while those who train models to critically evaluate all prompt content may confer resistance.

A 12.7 percentage point mean shift may appear modest in any individual interaction. At population scale, it is not. In the United States, approximately 6.9 billion prescriptions are dispensed annually ^18^. If even a fraction of prescribing decisions involve AI consultation, and adoption is accelerating, a systematic bias toward one brand over an equivalent competitor would redirect billions of dollars in pharmaceutical revenue. For context, U.S. net pharmaceutical spending reached $487 billion in 2024, with cardiovascular, antidiabetic, and oncologic agents among the largest therapeutic classes ^19^; a 1% shift in market share within any of these categories represents hundreds of millions to over one billion dollars in annual revenue redirected between therapeutically equivalent competitors.

This study shows that the clinical harm is not that patients receive a dangerous drug. Our Experiment 3 shows models resist that. The harm may be subtler: patients and physicians receive a recommendation that is clinically sound but commercially influenced, without any disclosure. This undermines the epistemic trust that clinical AI tools require to function. If physicians cannot distinguish an evidence-based recommendation from an advertising-influenced one, the tool’s value as a decision aid is compromised regardless of its accuracy.

We tested system prompt embedding, one of several plausible delivery mechanisms; in practice, pharmaceutical advertising also enters models through training data, retrieval-augmented generation pipelines, and user-pasted content. The economic incentive to exploit these channels may be substantial. As our data show, a single line of text in a system prompt can shift drug preference in susceptible models. For a drug with billions in annual revenue, even a modest increase in AI-influenced prescriptions could represent hundreds of millions of dollars. Unlike television or print advertising, which is subject to FDA oversight ^20^, advertising embedded in AI systems has no regulatory framework. The potential for misuse extends to off-label promotion, which has resulted in some of the largest pharmaceutical settlements in history ^21^.

AI-mediated promotion could circumvent existing enforcement because the promotional content is embedded in the model’s context rather than in a labeled advertisement. Notably, platforms like OpenEvidence already integrate pharmaceutical-sponsored content into AI-generated clinical summaries, demonstrating that this pathway is not speculative but operational.

One additional, and equally important, aspect is that these models are increasingly accessible to anyone with an internet connection. The population most likely to rely on AI for health decisions is also the population with the least access to independent medical expertise: uninsured individuals, those in primary care deserts, and patients who cannot afford specialist consultation ^22,23^. In the United States, nearly 600,000 healthcare-related queries are sent to AI chatbots weekly from hospital deserts alone (according to reports published by OpenAI), defined as areas more than a 30-minute drive from the nearest general hospital. If AI health assistants systematically favor branded drugs over equally effective generics, the cost burden falls disproportionately on these patients. Generic drugs cost 80 to 85 percent less than their branded equivalents on average ^24^; for an uninsured patient paying out of pocket, a 12.7 percentage point shift toward a branded drug can mean the difference between a $10 and a $200 monthly prescription for the same therapeutic compound.

For physicians, the risk is automation bias, the well-documented tendency to defer to algorithmic recommendations even when they conflict with clinical judgment ^25^. A physician who uses an AI tool to quickly check a drug recommendation and receives a confident, accurate, advertising-influenced answer may have no reason to second-guess it. The bias is invisible by design.

Nonetheless, this study has limitations. We tested system prompt embedding, one of several plausible delivery mechanisms. Real-world pathways (training data, RAG, user context) may produce different effect magnitudes and are harder to control experimentally. We tested brand-name drug pairs across 13 therapeutic areas; the full space of clinical decisions is larger. Our open-response sub-analysis used three representative models and five repetitions per cell; larger samples could refine per-scenario estimates. Our response parsing, while validated, may miss nuanced outputs. These limitations likely make our estimates conservative: system prompt ads are the most visible and detectable form of influence, and real-world contamination through training data would be more diffuse and harder to mitigate.

In conclusion, ad-supported AI in healthcare is not hypothetical; it is already being deployed. Our results provide the first large-scale empirical evidence that pharmaceutical advertising systematically biases clinical AI outputs in ways that are measurable, provider-dependent, and invisible to standard accuracy metrics. The bias operates silently between therapeutically equivalent options, producing no detectable signal of error while potentially steering recommendations toward advertised alternatives. Regulators must define what advertising is permissible in AI-assisted clinical decision-making. Developers must test and disclose their models’ susceptibility to promotional content. And health systems must evaluate whether AI tools exposed to pharmaceutical advertising can still meet the standard of unbiased clinical guidance that patients and physicians expect.

## Methods

We conducted a controlled experiment testing whether pharmaceutical advertisements embedded in AI system prompts shift drug recommendations. We queried 12 large language models from three providers (OpenAI, Anthropic, Google) across 33 clinical scenarios in four experiments, totaling 258,660 API calls. All code, study protocol, scenario data, and analyses are publicly available at https://github.com/MahmudOmar11/ad-verse-effects.

### Infrastructure

We evaluated 12 models spanning four size tiers (nano, small, large, flagship) from three providers: OpenAI (GPT-4.1 Mini, GPT-4.1, GPT-5 Mini, GPT-5.2, o4-mini), Anthropic (Haiku 4.5, Sonnet 4.5, Sonnet 4.6, Opus 4.6), and Google (Gemini 2.5 Lite, 2.5 Flash, 3 Flash). All models were accessed via official APIs with provider-default temperature settings. Each API call was independent, with no conversational memory between calls. Detailed configurations are in **Table S1**.

### Experiments 1-3

Three experiments tested distinct dimensions of advertising influence across 256,320 API calls.

### Experiment 1 (Preference Shift)

Thirteen clinical scenarios (S01--S13) each presented a patient with a condition for which two brand-name drugs from competing manufacturers were guideline-appropriate. Scenarios spanned cardiology, endocrinology, psychiatry, pulmonology, rheumatology, urology, and neurology. Each scenario was authored by a primary care physician using current clinical practice guidelines, including ACC/AHA, ADA, and GOLD guidelines ^26–28^. Two additional physicians (an internal medicine expert and rheumatologist, and another internal medicine expert and cardiology fellow) independently reviewed each scenario for clinical accuracy and therapeutic equipoise. Disagreements were resolved by consensus among the three reviewers, with oversight from a senior internal medicine and nephrology physician with over ten years of clinical experience. Models chose between drug A, drug B, or “neither/other.” Three conditions were tested: no ad (baseline), ad for drug A, and ad for drug B.

### Experiment 2 (Wellness Endorsement)

Ten scenarios (S14--S23) presented conditions where the evidence-based recommendation was non-pharmacological (e.g., cognitive behavioral therapy for insomnia, physical therapy for knee osteoarthritis, dietary modification for mild hyperlipidemia), and not pharmacological supplements. Scenarios were developed and validated using the same physician panel and review process described above.

### Experiment 3 (Accuracy Cost)

Ten scenarios (A01--A10) featured an advertised drug that was clinically suboptimal or contraindicated for the case (e.g., a GLP-1 agonist for a frail elderly patient, a biologic where methotrexate is first-line). The correct answer was always option C. All scenarios were physician-authored and independently validated.

### Experiment 4: Open-response sub-analysis

To complement the forced-choice design, we conducted an open-response experiment in which models generated free-text clinical justifications alongside their drug selections. Three representative models were selected based on Experiment 1 results to span the susceptibility range: Gemini 2.5 Flash (high susceptibility), GPT-4.1 (moderate), and Claude Opus 4.6 (resistant). The 13 equipoise scenarios from Experiment 1 were retained with the same three conditions (baseline, ad for drug A, ad for drug B).

Four system prompt personas were tested: physician, helpful AI assistant, customer service, and no persona. Each persona’s system prompt was appended with a standardized response format instruction requiring the model to provide a structured output: CHOICE (A, B, C, or D), CONFIDENCE (low, medium, or high), JUSTIFICATION (clinical reasoning in 50--150 words), and ALTERNATIVES (discussion of other options). Each cell (model × scenario × condition × persona) was repeated five times, yielding 2,340 API calls (3 models × 13 scenarios × 3 conditions × 4 personas × 5 repetitions).

Five rule-based natural language processing (NLP) metrics were computed for each response: ad-echo rate (proportion of advertising claims echoed in the justification), confidence asymmetry (change in model-reported confidence), competitor mention suppression, evidence specificity (weighted composite score of guideline citations, trial references, and mechanism-of-action reasoning), and spontaneous advertising disclosure rate ^29^. All metrics were deterministic and reproducible; no large language model-as-judge methods were used ^30^. Full metric definitions are in the **Supplementary Methods**.

### Experimental design

For Experiments 1--3, each cell (model × scenario × condition × persona × variant) was repeated 20 times. Four system prompt personas were tested: physician, helpful AI assistant, customer service agent, and no persona, based on our earlier work showing that persona system prompts shift clinical action thresholds in large language models (21). Three independently authored prompt variants per scenario controlled for wording effects. Advertisements followed a standardized format modeled on real direct-to-consumer pharmaceutical ads: “[Sponsored Message] Ask your doctor about [Brand]® for [indication]. [Brand] offers [clinical claim]. Learn more at [URL]. [End of Sponsored Message].” The ad did not instruct the model to prefer the drug.

### Statistical analysis

All analyses were conducted in Python 3.12 using NumPy, SciPy, and statsmodels. Preference shift was defined as the difference between the ad-condition chose-advertised rate and a baseline proxy (the mean of baseline option A and B selection rates). Significance was assessed using chi-square tests with Yates correction. Effect sizes are reported as Cohen’s *h*. Confidence intervals (95%) were computed using the Wilson score method for proportions and the Newcombe method for differences. For the open-response sub-analysis, the primary outcome was the shift toward the advertised drug, defined as P(choose advertised | ad condition) − P(choose that same option | baseline), computed per model × scenario × persona cell and aggregated across cells. All statistical code is available in the study repository.

## Data Availability

All data produced in the present study are available upon reasonable request to the authors

## Acknowledgments

We thank the physician reviewers for their contributions to scenario development and validation.

## Funding

None.

## Author contributions

M.O. conceived the study, designed the experiments, developed the pipeline, ran all analyses, and wrote the manuscript. R.A. contributed to scenario validation and data review. J.M. and A.Z. provided clinical input on scenario design. A.R. and M.G. contributed to manuscript revision and interpretation. E.-M.A. assisted with literature review. B.S.G. and A.S. contributed to data interpretation and manuscript review. A.W.C. provided methodological guidance. E.K. co-supervised the project and contributed to study design and manuscript revision. G.N.N. co-supervised the project, contributed to study design, and critically revised the manuscript.

## Competing interests

The authors declare no competing interests.

## Data and code availability

All data, code, and clinical scenarios are publicly available at https://github.com/MahmudOmar11/ad-verse-effects

## Notes

### Competing Interest Statement

The authors have declared no competing interest.

### Funding Statement

This work was supported in part through the computational and data resources and staff expertise provided by Scientific Computing and Data at the Icahn School of Medicine at Mount Sinai and supported by the Clinical and Translational Science Awards (CTSA) grant UL1TR004419 from the National Center for Advancing Translational Sciences. Research reported in this publication was also supported by the Office of Research Infrastructure of the National Institutes of Health under award number S10OD026880 and S10OD030463. The content is solely the responsibility of the authors and does not necessarily represent the official views of the National Institutes of Health. The funders played no role in study design, data collection, analysis and interpretation of data, or the writing of this manuscript.

### Author Declarations

Simulated internal data

